# Comorbidities & coronary vascular disease (CVD) conditions in impaired cognitive disease with coronary artery disease (ICAD) in *AllofUS*

**DOI:** 10.1101/2025.04.30.25326756

**Authors:** Praveen Hariharan, Minoo Bagheri, Emelia Asamoah, Ioana Voiculescu, Purnima Singh, Tafadzwa Machipisa, Tess Pottinger, Antone Opekun

**Author notes:** Corresponding Author: Praveen Hariharan, Department of Emergency, Brown University Health, Warren Alpert Medical School, 223 walnut st, #20, Framingham, MA.

## Abstract

**Background and objectives:** Impaired cognitive disease (IC) as a clinical entity may have heterogeneous neuropathologies and share similar risk factors with coronary artery disease (CAD). To evaluate the association of cardiovascular disease (CVD) conditions and comorbidities in IC with coronary artery disease (ICAD) compared to CAD alone and determine the effect modification when CAD preceded (ICAD1) or succeeded Impaired cognitive disease (ICAD2) event in ICAD patients.

**Design and setting:** We included unique *AllofUS* participants (aged >60 yrs) with IC (n=6606), CAD (n=27516), ICAD (n=4852), and healthy controls (n=78,600) and used validated ICD-9/10 codes for defining exposures. We performed multivariable logistic regression of CVD conditions (ventricular fibrillation, cardiac arrest, conduction deficits, pacemaker, cardioversion, diastolic and systolic dysfunction) and comorbidities, in particular, CABG, in ICAD compared to CAD alone after adjusting for traditional risks (age, sex, depression, diabetes, hypertension, smoking, alcohol use, and stroke). We performed sensitivity analyses after propensity matching, restricting only outpatient confirmed diagnoses, those with a history of left heart catheterization (LHC), and CABG. We performed similar stratified analyses in ICAD1 and ICAD2 based on CAD preceding or succeeding IC events by at least one year. We defined significance at p <0.05.

**Results:** CVD conditions, such as ventricular fibrillation (1.38;1.09-1.74, p=0.007), atrioventricular blocks (1.15;1.04-1.27, p=0.005), pacemaker (1.37;1.10-1.69, p=0.003), cardioversion (1.65;1.12-2.39, p=0.009), and diastolic dysfunction (1.09;1.01- 1.18, p=0.03) were associated with ICAD compared to CAD alone with higher associations in LHC and CABG patients. CVD conditions were significantly higher in ICAD1 than in ICAD2 compared to CAD. In the adjusted multivariate model, CABG (1.44;1.25-1.64, p<0.001) was associated with ICAD1.

**Conclusions:** CVD conditions and CABG were significantly associated with ICAD when CAD preceded IC. Accounting for these comorbidities and CVD complications while assessing CAD patients can help in clinical risk stratification for ICAD.

**Lay summary:**

1. We used NIH-AllofUS (n=117,574) participants (aged >60 yrs) to assess how specific cardiovascular disease (CVD) conditions (ventricular fibrillation, cardiac arrest, conduction deficits, pacemaker, cardioversion, diastolic dysfunction) and comorbidities were associated with age-related impaired cognitive disease (IC) with CAD (ICAD) compared to CAD alone and if that association changes when CAD preceded IC event in ICAD.
2. We found that CVD conditions were associated with ICAD compared to CAD (without IC). Comorbidities, particularly coronary artery bypass grafting, were also significantly associated with ICAD when CAD preceded an IC event.
3. The association of CVD conditions and CABG with ICAD suggests a more clinically severe CAD phenotype in ICAD when the CAD event precedes IC.

## INTRODUCTION

Coronary Artery Disease (CAD) and Impaired Cognitive Disease (IC), in particular Alzheimer’s Disease and related dementias (ADRD), are a significant cause of morbidity in the US and globally. ^1–3^ Traditional risk factors like age, sex, diabetes, hypertension, stroke, depression, smoking, and alcohol use are shared by CAD and IC patients. ^4,5^

IC in CAD (ICAD) prevalence increases with age in geriatric patients (>60 years), yet risk stratification for ICAD in CAD patients remains suboptimal.^2,6^ Many individual studies have identified unique comorbidities related to mental health, neurologic conditions, cardiac comorbidities, endocrinopathy, bleeding disorders, micronutrient deficiency, and renal dysfunction to be associated with IC or geriatric CAD patients separately. ^4,7–14^ However, the association of such comorbidities with ICAD compared to CAD alone is largely unknown. In addition, coronary artery bypass grafting (CABG) as a comorbidity has been associated with short-term cognitive decline, but large studies assessing its association with ICAD are lacking.^15^ Understanding the demographics and burden of comorbidities can lead to heightened evaluation of CAD patients for better risk stratification of ICAD and provide insight into the clinical pathophysiology of ICAD.

ICAD has higher age-adjusted mortality compared to IC and CAD alone.^2^ However, the burden of cardiovascular disease (CVD) conditions (ventricular fibrillation, cardiac arrest, conduction deficits, pacemaker, cardioversion, and systolic/diastolic dysfunction) in ICAD compared to CAD without IC is unclear.^2^ It is also unclear if this association of comorbidities and CVD conditions with ICAD is modified when CAD precedes IC in ICAD. In this context, we sought to evaluate the association of comorbidities and CVD conditions with ICAD compared to CAD without IC. We also investigated the effect modification when CAD preceded (ICAD1) or succeeded the IC event (ICAD2) in ICAD patients using the NIH *AllofUS* clinical cohort.

## METHODS

### a) Study Design

eFigure 1 in Supplement 1 highlights the case-control study design. We report our study according to STROBE guidelines.

### b) Setting and Data Sources

We used the NIH *AllofUS* clinical registered tier V7 dataset made available to authorized users on the researcher workbench for our analyses. In brief, the *AllofUS* research program assembles participant data from diverse participants across 50 healthcare organizations in the US and includes surveys, electronic health records (EHR), biosamples, and genomic data. ^16^ *AllofUS* utilizes the Observational Medical Outcomes Partnership Common Data Model (OMOP CDM) to standardize data. ^16^

### c) Selection of Cases and Controls

We used OMOP CDM v 5.3.1 concept codes based on ICD-9/10 and CPT (Current Procedural Terminology) codes to define our phenotypes (IC, CAD, & ICAD), comorbidities, CVD conditions, and used associated index date of diagnoses.^5,11,17–20^

#### IC definition

We have defined IC based on the ICD-9/10 diagnosis corresponding to dementia (290.X, 291.X, 294.X, 331.X, F00.X, F00.X, F01.X, F02.X, F03.X, F10.X, G30.X, G31.X), or impaired memory (R41.3, 780.93).^5,11,18,20^

#### CAD definition

Any patient meeting one of the following criteria: A history of myocardial infarction (MI) (ICD-9: 410.x-412.x;ICD- 10: I21.X-I24.X); the presence of stable or unstable angina (ICD-9: 411.x-413.x;ICD-10: I20.X); a history of chronic ischemic heart disease (ICD-9:414.x; ICD-10: I25.X); a history of percutaneous coronary intervention (PCI, ICD-10: Z95.5, Z98.61) or CABG (ICD-10: Z95.1). ^5,19,21,22^

#### ICAD definition

We defined impaired cognitive disease with coronary artery disease (ICAD) when patients have both IC and CAD. Based on the index diagnosis date, we further defined ICAD1 when CAD preceded IC and ICAD2 when CAD succeeded IC event by at least one year. We determined a window of one year to account for potential temporal delays in EHR diagnoses date recording by health care providers.^23^

#### Healthy controls

We defined healthy controls as patients not meeting the IC or CAD definition. In addition, healthy controls lacked any other OMOP concept codes corresponding to hypertension, diabetes, kidney disease, hyperlipidemia, late effects of cerebrovascular disease, cardiac arrhythmia, or obesity.

### d) Comorbid Variables

We have defined our comorbid variables based on previously published reports of association with IC or geriatric CAD corresponding to their respective OMOP concept codes ^4,5,7,10,13,14^:

a. <u>Comorbidities:</u>
  1) Mental health: Anxiety, depression, and psychosis.
  1) Neurologic conditions: Ischemic stroke, transient ischemic attack, intracranial hemorrhage (ICH), seizure disorder, and dysosmia/dysgeusia.
  2) Cardiac comorbidities: Syncope, hypertension, hypertensive urgency, mitral regurgitation, obstructive sleep apnea (OSA), atrial fibrillation (AF), CABG, and PCI.
  3) Endocrinopathy: Osteoporosis and hypothyroidism.
  4) Bleeding conditions: Upper gastrointestinal bleeding (UGIB) and lower gastrointestinal bleeding (LGIB).
  5) Micronutrient deficiency: Iron deficiency, and vitamin B12 deficiency.
  6) Renal dysfunction: Chronic kidney disease (CKD), dialysis, and proteinuria.
b. <u>CVD conditions:</u> Ventricular fibrillation (VF), cardiac arrest (CA, includes VF), atrioventricular blocks (AVB), pacemaker implant (PPM), cardioversion, diastolic dysfunction, and systolic dysfunction.^5,10^

### e) Statistical Analysis

#### 1) Primary Analyses (e Methods in Supplement 1)

a. For our primary analyses, we included participants with at least one hospital or outpatient visit encounter meeting our phenotypic definition who were older than 60. We analyzed baseline characteristics with simple means and proportions. Comorbidities demonstrating suggestive differences across phenotypes with p≤0.1 in univariate analysis were selected for all multivariable logistic regression analyses After adjusting for traditional risks, we finally retained clinically meaningful predictor variables for the final regression model.
b. To assess effect modification based on CAD temporality to IC event in ICAD, we performed stratified analysis in ICAD1 and ICAD2.

#### 2) Sensitivity and Subgroup Analyses (e Methods in Supplement 1)

We performed sensitivity analyses to ensure consistency of our findings:

a. Propensity Score Matched Analyses: Given that the traditional risk factors in ICAD can confound the association of some comorbidities and CVD conditions between ICAD and CAD, we propensity-matched the ICAD group to CAD for all traditional risks. We used a logistic regression model with nearest neighborhood matching, maintaining a treated-control ratio 1:3. ^10,24,25^
b. Outpatient Diagnosis Analyses: To increase the accuracy of phenotype definitions primarily related to IC, we performed a sensitivity analysis by including IC, CAD, or ICAD cases diagnosed only in the outpatient setting due to the possibility of accuracy gaps in IC-related billing codes in the inpatient setting.^18,26^
c. Left heart catheterization (LHC) and CABG patients: To increase the accuracy of CAD definition and understand the role of CABG, we performed a sensitivity analysis by including CAD, ICAD patients with history of left heart catheterization and CABG diagnosed through validated CPT codes.^19^

#### 3) Statistical tools (e Methods in Supplement 1)

We performed all the analyses in R integrated within Jupyter Notebooks (a secure, access-controlled, and cloud-based analysis environment) in the AllofUS researcher workbench. We used the *matchit* and samplesizelogisticcasecontrol packages for propensity-score matching and sample size estimation, respectively.^27,28^ Based on prior reports and our data, we used 10 percent prevalence of ICAD in CAD. ^2,5,29^ For primary analyses, by enrolling ~ 32,000 patients (case: controls 1:5), we estimated to detect a minimum odds ratio (OR) for binary exposures of up to 1.20, at 80% power, with a minimum level exposure in controls at 3.5%. We defined the statistical significance of P values for all analyses at 2-sided P□values <□0.05.

## RESULTS

We screened 134,663 patients with at least one hospital or outpatient encounter, meeting our phenotypic definitions (eFigure 1 in Supplement 1). We excluded 640 with mismatched ICD-9/10 codes and 16,449 with age <= 60 years. Finally, we included 117,574 patients (6,606 with IC, 27,516 with CAD, 4,852 with ICAD, and 78,600 healthy controls) above 60 years old.

### A) Primary Analyses

In the univariate analyses (Table 1), we noted ICAD patients were significantly (p<0.001) older (75.5 years, SD=7.5) compared to CAD (73.5 years, SD=7.4), IC (73.3 years, SD=7.4), and healthy controls (70.8 years, SD=6.9). The proportion of females was highest in IC (62%), followed by healthy controls (55%), ICAD (49%), and CAD (44%, p<0.001). ICAD patients had a significantly higher proportion of comorbidities compared to the other three groups (Table 1).

**Table 1:** Univariate analysis (*AllofUS, Age >60*)

| Predictors | Controls<br>N=78,600 | IC<br>N=6,606 | CAD<br>N=27,516 | ICAD<br>N=4,852 |
| --- | --- | --- | --- | --- |
| **Demographics\*** |  |  |  |  |
| Age, mean (SD) | 70.8 (6.9) | 73.3 (7.4) | 73.5 (7.4) | **75.5 (7.5)** |
| Female Sex | 43,406 (55.2%) | 4,109 (62.2%) | 12,100 (44%) | **2,383 (49.1%)** |
| **Race\*** |  |  |  |  |
| White | 51,138 (65.1%) | 4,511 (68.3%) | 17,955 (65.3%) | **3,148 (64.9%)** |
| Black | 13,601 (17.3%) | 814 (12.3%) | 4,650 (16.9%) | **722 (14.9%)** |
| **Ethnicity\*** |  |  |  |  |
| Hispanic | 8,002 (10.2%) | 863 (13.1%) | 3,107 (11.3%) | **632 (13.0%)** |
| **Mental Health Comorbidities\*** |  |  |  |  |
| Depression | 1,762 (2.2%) | 3,631 (55%) | 10,452 (38%) | **3,149 (64.9%)** |
| Anxiety | 1,629 (2.1%) | 3,172 (48%) | 9,688 (35.2%) | **2,747 (56.6%)** |
| Psychosis | 231 (0.3%) | 429 (6.5%) | 1,082 (3.9%) | **489 (10.1%)** |
| **Neurologic Comorbidities\*** |  |  |  |  |
| Intracranial Hemorrhage | 0 | 254 (3.8%) | 946 (3.4%) | **410 (8.5%)** |
| Ischemic Stroke | 69 (0.1%) | 646 (9.8%) | 2,887 (10.5%) | **1101 (22.7%)** |
| Transient Ischemic Attack | 0 | 588 (8.9%) | 2,169 (7.9%) | **886 (18.3%)** |
| Seizure | 165 (0.2%) | 466 (7.1%) | 1,136 (4.1%) | **505 (10.4%)** |
| Dysosmia/Dysgeusia | 71 (0.1%) | 157 (2.4%) | 329 (1.2%) | **133 (2.7%)** |
| **Cardiac Comorbidities\*** |  |  |  |  |
| Syncope | 225 (0.3%) | 669 (10.1%) | 2,966 (10.8%) | **1,015 (20.9%)** |
| Hypertension | 0 | 1,198 (18.1%) | 6,676 (24.3%) | **1,650 (34.0%)** |
| Hypertensive Urgency | 0 | 217 (3.3%) | 2,047 (7.4%) | **491 (10.1%)** |
| Mitral Regurgitation | 205 (0.3%) | 359 (5.4%) | 3,794 (13.8%) | **765 (15.8%)** |
| Obstructive Sleep Apnea | 15 (0.7%) | 2,118 (32.1%) | 8,956 (32.5%) | **2,260 (46.6%)** |
| Atrial Fibrillation | 379 (0.5%) | 650 (9.8%) | 7,651 (27.8%) | **1,545 (31.8%)** |
| CABG | 0 | 0 | 1767 (6.4%) | **384 (7.9%)** |
| LHC | 0 | 0 | 6101 (22.2%) | **1190 (24.5%)** |
| PCI | 0 | 0 | 1651 (6%) | **331 (6.8%)** |
| **Endocrinopathy\*** |  |  |  |  |
| Type 2 Diabetes | 0 | 1,934 (29.3%) | 12,388 (45.0%) | **2,508 (51.7%)** |
| Hypothyroidism | 903 (1.1%) | 1,381 (20.9%) | 4,858 (17.7%) | **1,273 (26.2%)** |
| Osteoporosis | 1477 (1.9%) | 1,764 (26.7%) | 5,252 (19.1%) | **1478 (30.5%)** |
| **Bleeding comorbidities\*** |  |  |  |  |
| Upper GI Bleeding | 68 (0.1%) | 165 (2.5%) | 1,415 (5.1%) | **384 (7.9%)** |
| Lower GI Bleeding | 371 (0.5%) | 757 (11.5%) | 3,489 (12.7%) | **940 (19.4%)** |
| **Micronutrient deficiency\*** |  |  |  |  |
| Iron deficiency | 436 (0.6%) | 1,063 (16.1%) | 5,552 (20.2%) | **1,444 (29.8%)** |
| B12 deficiency | 80 (0.1%) | 307 (4.6%) | 1,053 (3.8%) | **347 (7.2%)** |
| **Kidney Disease\*** |  |  |  |  |
| Chronic Kidney Disease | 0 | 1,024 (15.5%) | 8,384 (30.5%) | **1,824 (37.6%)** |
| Proteinuria | 46 (0.1%) | 338 (5.1%) | 2,463 (9%) | **608 (12.5%)** |
| Dialysis | 2 (0%) | 27 (0.4%) | 859 (3.1%) | **132 (2.7%)** |
| **Lifestyle\*** |  |  |  |  |
| Alcohol Use | 590 (0.8%) | 627 (9.5%) | 2,739 (10.0%) | **650 (13.4%)** |
| Smoking | 1,250 (1.6%) | 1,045 (15.8%) | 7,063 (25.7%) | **1,343 (27.7%)** |
| Opioid Use | 194 (0.2%) | 185 (2.8%) | 1,160 (4.2%) | **286 (5.9%)** |

\*Proportions in bold with P value <0.001 (Pearson's Chi-squared test, baseline group = Controls)
CAD: Coronary Artery Disease without IC; ICAD: Impaired Cognitive Disease with Coronary Artery Disease; IC: Impaired Cognitive Disease without CAD
GI: Gastrointestinal; CABG: Coronary artery bypass graft; LHC: Left heart catheterization; PCI: Percutaneous coronary intervention

In our stratified analysis (eTable 1a/1b), while both ICAD1 (47%) and ICAD2 (55%) had a significantly higher proportion of females (p <0.05) compared to CAD (44%), ICAD1 had higher proportion of comorbidities like hypertension (40% vs. 27%, p<0.001), type 2 diabetes (54 % vs. 46%, p<0.001), CKD (40% vs. 32 %, p<0.001), AF (35% vs. 26%, p <0.001), CABG (11 % vs. 2.1%, p <0.001), LHC (29% vs 17.2%, p<0.001) and smoking (29% vs. 25%, p=0.009) compared to ICAD2 (eTable 1a/1b in Supplement 1). ICAD females had a lower proportion of LHC (21% vs. 28%, p <0.001) and CABG (4% vs. 12%, p<0.001) compared to ICAD males.

When we compared CVD conditions across all groups (Table 2), ICAD had a significantly higher proportion of CVD conditions (VF, CA, AVB, PPM, cardioversion, diastolic dysfunction, and systolic dysfunction) than CAD, IC, and healthy controls (Table 2). Most of the CVD conditions preceded the onset of IC in ICAD. The proportion of CVD conditions was even higher in ICAD when we restricted our analysis of LHC and CABG patients (Table 2).

**Table 2:**
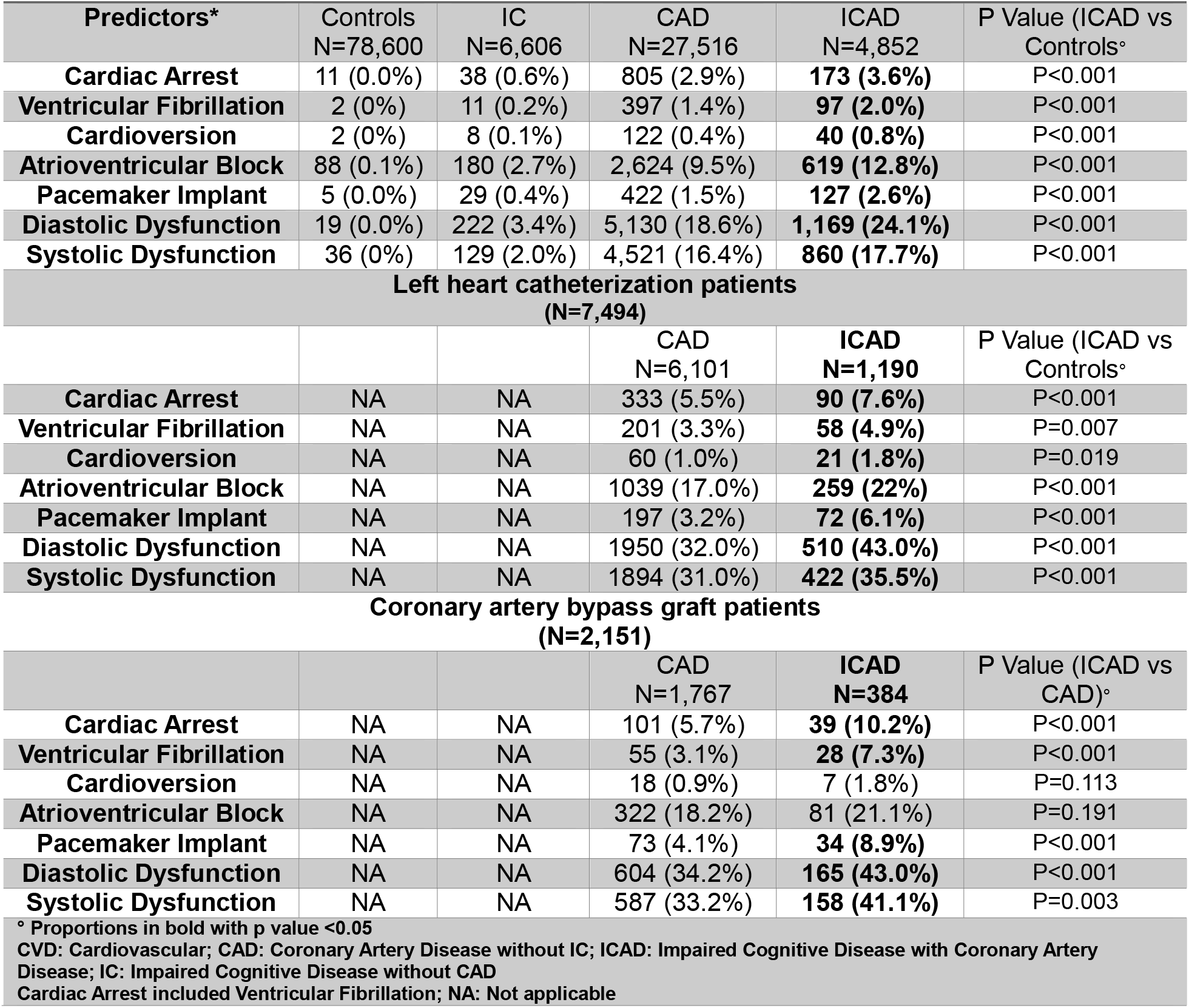
CVD conditions (*AllofUS, Age >60)*

In adjusted multivariable logistic regression of comorbidities (Figure 1), ICAD patients had higher odds of multiple comorbidities than CAD patients. These associations persisted in the stratified analysis (ICAD1/ICAD2 vs. CAD). In particular, we noted the association of CABG with ICAD1 (1.44;1.25-1.63, p <0.001) (Figure 2). We also noted an incremental association for each year of CABG with ICAD1 compared to CAD (1.03; 1.02-1.04, p <0.001). When we compared IC patients to CAD patients, only specific comorbidities related to mental health conditions, neurologic conditions, and endocrinopathy (osteoporosis) had significantly higher associations with IC patients (eFigure 2 in Supplement 1).

**Figure 1:**
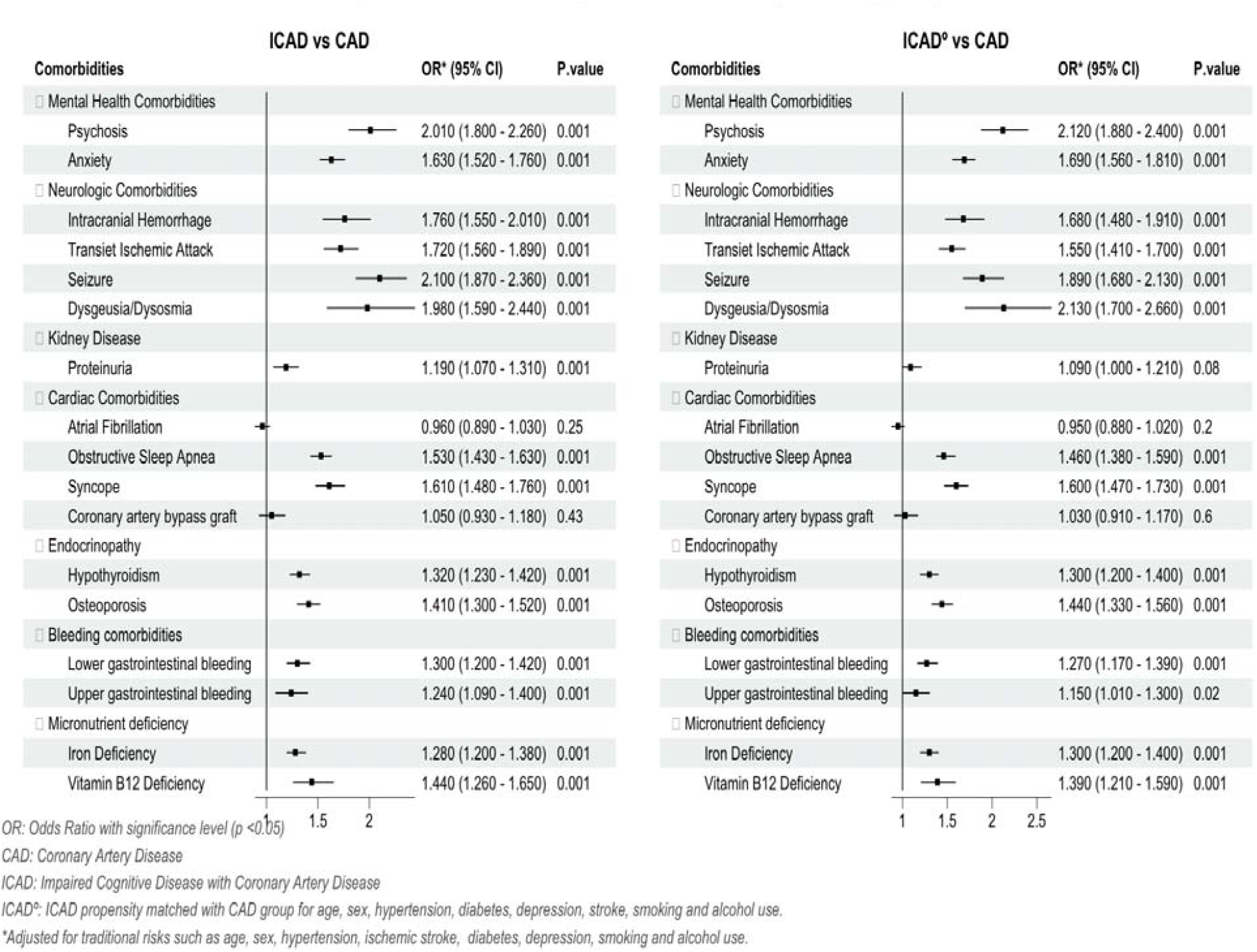
Multivariable analyses of comorbidities (AlloflJS, Age >60)

**Figure 2:**
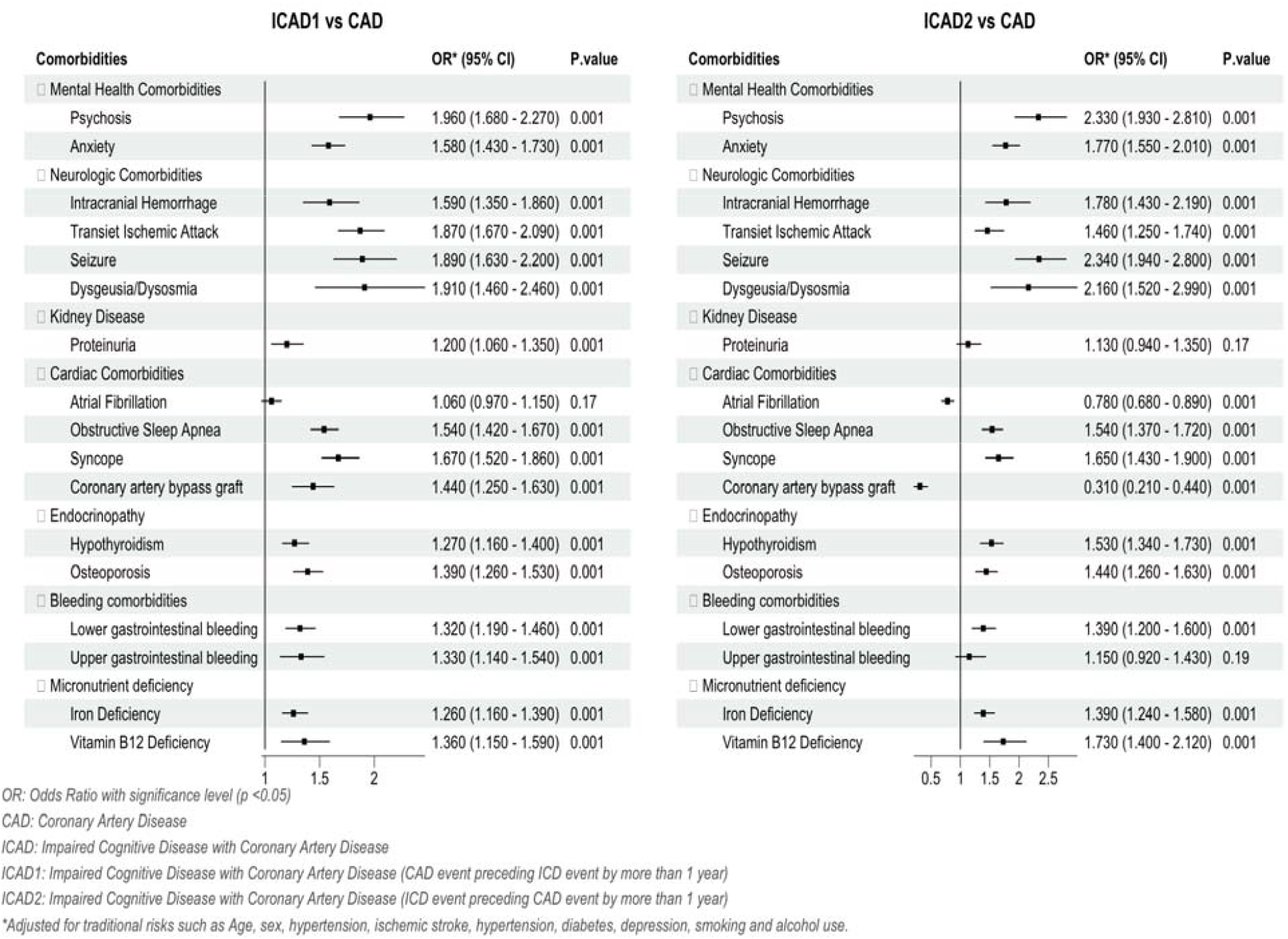
Multivariable analyses of comorbidities (AllofUS, Age >60)

In adjusted multivariable analysis of CVD conditions (Figure 3), ICAD had higher odds of certain CVD conditions such as VF (1.38;1.09-1.74, p=0.007), AVB (1.15; 1.04-1.27, p=0.005), PPM, (1.37;1.10-1.69, p=0.003), cardioversion (1.65; 1.12- 2.39, p=0.009), and diastolic dysfunction (1.09; 1.01-1.18, p=0.03). All CVD conditions had a significant association with ICAD1 but were non-significant with ICAD2 compared to CAD (Figure 3). In addition, we observed no association of systolic dysfunction with ICAD (0.96; 0.88-1.05, p=0.44) and lower odds in ICAD2 (0.63; 0.52-0.75, p<0.001) compared to CAD (Figure 3).

**Figure 3:**
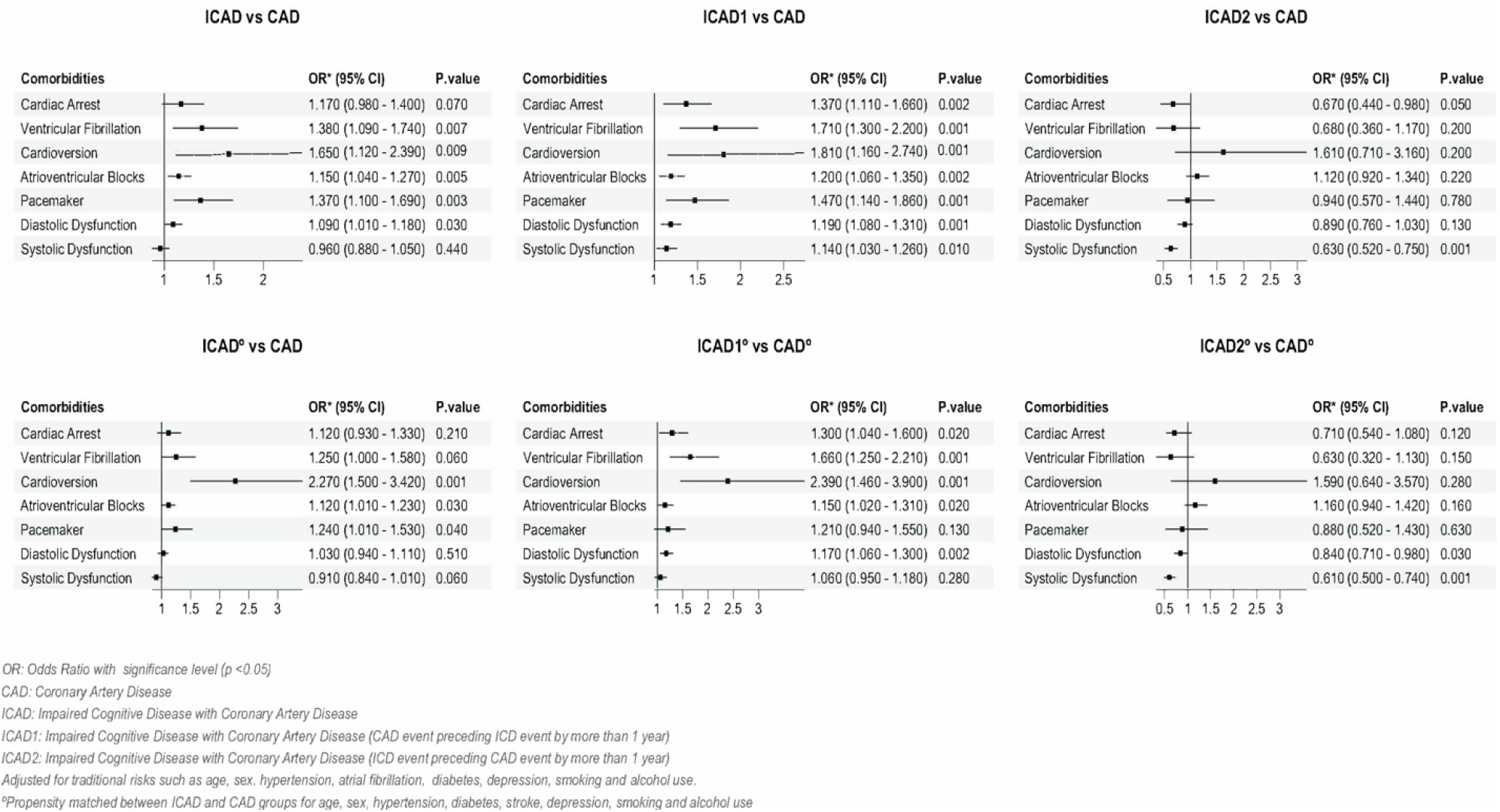
Multivariable analyses of cardiovascular conditions (AllofUS, Age >60)

### B) Propensity Score Matched Analyses

After propensity-score matching between ICAD and CAD, we noted a similar association of comorbidities (eFigure 3 in Supplement 1) and CVD conditions with ICAD and ICAD1/ICAD2 compared to CAD except mild attenuation of the association between VF and diastolic dysfunction with ICAD compared to CAD (Figure 1, Figure3).

### C) Outpatient Diagnosis Analyses

When we restricted our phenotypic definitions to the outpatient settings, we observed a similar distribution of demographics and comorbidities across phenotypes (eTable 2 & eTable 3a/3b in Supplement 1) and a similar association of comorbidities with ICAD and ICAD1/ICAD2 compared to CAD (eFigure 4 & eFigure 5 in Supplement 1). The proportion of CVD conditions was higher in the ICAD group than in other groups (eTable 4a in Supplement 1). In adjusted multivariable analyses of CVD conditions, we noted mild attenuation of association of certain CVD conditions (CA, VF, PPM, diastolic dysfunction) with ICAD while, AVB remained significantly associated with ICAD and ICAD1. (eFigure 6 in Supplement 1).

### D) LHC and CABG patients

We noted a stronger association of all CVD conditions with ICAD and ICAD1 among LHC patients compared to CAD (e Figure 7 in Supplement 1). The adjusted odds for VF and CA were significantly higher in ICAD at 1.52 (1.10-2.08, p=0.008) and 1.40 (1.08-1.80, p=0.01), respectively. The adjusted odds for PPM and diastolic dysfunction were 1.57 (1.16-2.10, p=0.003) and 1.28 (1.10-1.50, p<0.001) respectively (e Figure 7 in Supplement 1). We observed similar results in the CABG patients (data not shown).

## DISCUSSION

In our large case-control risk factor analyses of AllofUS participants (age > 60 yrs), we observed an increased proportion of CVD conditions (VF, AVB, PPM, cardioversion, and diastolic dysfunction) with ICAD compared to CAD without IC after adjusting for traditional risks. CVD conditions were more significantly associated with ICAD in the left heart catheterization group and CABG group, suggestive of more severe CAD phenotype in ICAD. Most CVD conditions (CA, VF, AVB, PPM, cardioversion, and diastolic dysfunction) had a significant association with ICAD when CAD preceded IC event, i.e., ICAD1. We also observed many co-morbidities related to mental health neurologic conditions, renal dysfunction, cardiac comorbidities, endocrinopathy, bleeding conditions, and micronutrient deficiency to be associated with ICAD compared to CAD without IC. The association of comorbidities persisted regardless of the temporality of CAD events around IC events in ICAD. Accounting for these comorbidities and CVD complications while assessing CAD patients can help in clinical risk stratification for ICAD

Cognitive decline after a CAD event is a matter of concern.^5,6,30^ However, risk stratification for IC remains suboptimal in CAD patients. We noted increased odds of almost all CVD conditions with ICAD except systolic dysfunction (Figure 3). In our stratified analyses, we observed effect modification, where all CVD conditions were significantly higher in ICAD1 than in ICAD2 (Figure 3) that increased LHC patients (eFigure 7 in Supplement 1). In particular, we observed higher OR for arrhythmic conditions like VF, CA, AVB, and PPM in ICAD1. The association of pacemaker implants with vascular dementia has been previously described in a large Danish cohort of MI patients.^5^ In addition, sex differences between ICAD2 and ICAD1 (females;55% vs 49%, p<0.001) could play an essential role in the effect modification. While we noted lower revascularization rates in females across all groups, ICAD males received more revascularizations compared to CAD males. A study evaluating dementia patients with a history of MI noted lower comorbidities in females compared to males, and females received lesser invasive management of MI.^31^ Also, given that CAD and IC have polygenic influence, underlying genetic architecture differences could contribute to the differences in CVD conditions between ICAD1 and ICAD2. ^21,32^

Coronary artery bypass graft (CABG) has been associated with cognitive decline.^15^ However, studies evaluating the association of CVD conditions in CABG with ICAD are lacking.^15^ In our study, CABG, along with the duration of CABG, was associated with ICAD1 as compared to ICAD2 in all analyses, including in the angiography subset, suggestive of increased vulnerability in CABG patients.^15^ A recent meta-analysis failed to identify a single risk factor associated with long-term IC after CABG due to lack of power, lack of follow-up, and heterogeneity between individual studies.^15^ While our study did not evaluate the association of the anatomy of CABG with ICAD1, it seems consistent with a recent study by Olesen et al., which noted a higher risk of ADRD in 3-vessel disease as compared to 1 or 2-vessel disease.^29^ Using our large sample size (CABG, n=2,151), we identified several CVD conditions associated with ICAD. However, the mechanisms linking comorbidities and CVD conditions with ICAD in patients undergoing CABG need further investigation to identify risk reduction strategies.

Diastolic dysfunction has been associated with AD/ADRD, possibly through the deposition of amyloid precursor protein Aβ and phosphorylated tau protein in the myocardium based on autopsy studies and animal models.^33,34^ In our adjusted models, ICAD had higher odds of diastolic dysfunction than CAD patients (without IC), more significant in ICAD1 and when we restricted to LHC and CABG cases (Table 2, e Table 6 in Supplement 1). The association of systolic dysfunction with IC is inconsistent. ^35^ It has been speculated chronic cerebral hypoperfusion from cardiac dysfunction is a risk factor for ICAD. However, we did not observe any association of systolic dysfunction with ICAD in our primary or sensitivity analyses. ^6,35,36^ Our findings are consistent with a recent study by Huynh et al., which reported a lack of association between cognitive impairment and systolic dysfunction. ^37^ However, investigating the role of diastolic dysfunction linking CVD conditions in ICAD when CAD precedes IC event is further warranted.

## Limitations

One of the primary challenges of the case-control study is retrospective nature and the potential for information bias and measurement bias. We defined CAD and IC based on previously validated ICD-9/10 algorithms using EHR data. ^11,19–21^. We acknowledge the challenges in accuracy for identifying IC solely based on a set of ICD-9/10 codes. However, we tried to increase the likelihood of true diagnosis by restricting to age above 60 years and by performing a sensitivity analyses using outpatient-only diagnosis. ^18,26^ We used an EHR-based index date of diagnosis for our phenotypes and acknowledged that there could be some discordance between the “true onset of disease” and EHR recorded diagnosis date.^23^ While we had adequate power (>80%) for our primary analyses, we did not have enough power for all subgroup analyses. We also acknowledge the possibility of unmeasured confounders affecting our association analyses, but we tried to limit its extent by using propensity score matching of traditional risks.^10,24,25,27^ However, our case-control study design using the *AllofUS* dataset with a standardized data collection model that integrates decades of EHR data for each patient can somewhat overcome these biases and provide a stepping stone for future prospective studies in this field.^38^

We identified many comorbidities significantly associated with ICAD compared to CAD, regardless of CAD temporality. However, CVD conditions and CABG were significantly associated with ICAD when CAD preceded IC. The association of CVD conditions and CABG with ICAD suggests a more severe clinical CAD phenotype in ICAD.yw

## Data Availability

All data produced in the present work are contained in the manuscript.

## Acknowledgments

We gratefully acknowledge *AllofUs* participants for their contributions, without whom this research would not have been possible. We also thank the NIH *AllofUs* Research Program for making available the participant data examined in this study.

## Author Contributions

The team had full access to all of the data in the study and take responsibility for the integrity of the data and the accuracy of the data analysis.

Concept and Design: All authors

Acquisition, analysis, or interpretation of data: Dr.Hariharan, Dr. Asamoah, and Dr.Bagheri

Drafting of the initial manuscript: Dr.Hariharan

Statistical analysis: Dr.Hariharan, Dr.Bagheri, Dr.Pottinger, and Dr.Machipisa.

Critical review of the manuscript for important intellectual content: Dr.Singh, Dr.Voiculescu, Prof A Opekun, Dr.Josee Dupuis, and Dr.J Dawn Abbott.

## Disclaimer

The views expressed are those of the authors and not necessarily those of the funders, NIH, or *AllofUS* research program.

